# Pathogenicity evaluation of coding germline variants identifies rare alleles enriched in hematological patients of a founder population

**DOI:** 10.1101/2024.10.23.24315723

**Authors:** Jessica R. Koski, Laura Langohr, Tuulia Räisänen, Atte K. Lahtinen, Marja Hakkarainen, Caroline A. Heckman, Ulla Wartiovaara-Kautto, Esa Pitkänen, Outi Kilpivaara

## Abstract

**Background:** The clinical significance of most germline variants in hematological malignancies (HMs) remains unknown. This presents a challenge in the clinical setting, as the inability to accurately detect pathogenic variants can influence therapeutic decisions. Population isolates have been shown to be beneficial in pathogenic variant discovery due to presence of rare deleterious variants in relatively high frequencies.

**Methods:** We developed and applied PaVaDi, a computational pipeline that follows American College of Medical Genetics and Genomics (ACMG) guidelines, to evaluate the pathogenicity of germline variants in 511 HM patients from the Finnish founder population. We conducted an exome-wide burden analysis to assess the overall contribution of pathogenic variants to HMs and identified significant gene associations. We also examined genes previously associated with hematological diseases and DNA repair in more detail, and performed protein stability analyses to resolve variants of unknown significance (VUS).

**Results:** The exome-wide burden analysis revealed potential pathogenic alleles in *CUX2, RNPC3,* and *MFSD2A* that have not previously been linked to HM predisposition. We also identified the largest series of *CHEK2* variant carriers reported in hematological diseases, including pathogenic/likely pathogenic (P/LP) variants (*n*=19), Ile200Thr (*i.e.*, Ile157Thr) (*n*=49), and other variants of uncertain significance (*n*=3). *CHEK2* variants were 1.7-fold enriched in patients compared to controls (13.9% vs 8.3%, *p*=2×10^−5^). Strikingly, Ile200Thr was enriched over four-fold in acute lymphoblastic leukemia patients. Finally, protein structure stability analyses suggested novel *MPO* variants to be potentially highly deleterious.

**Conclusions:** This study highlights the importance of germline testing in hematological malignancies and demonstrates the utility of population isolates for pathogenic variant discovery. Our findings identify a significant burden of deleterious variants in HM patients, particularly in *CHEK2*, and underscore the potential of multi-disease joint analyses in revealing germline contributions to hematological diseases.

## Background

Interpretation of germline genetic analyses challenges modern hematological disease diagnostics and care. In clinical practice, the quality of pathogenicity evaluation can vary substantially depending on evaluation procedures in place and training of clinical geneticists (1,2). Guidance established by ACMG is being used to combine evidence supporting variant pathogenicity, providing a rule-based framework to classify germline variant pathogenicity into five categories (3). The ACMG framework integrates functional evidence, *in silico* pathogenicity predictions, population allele frequency information and other sources of evidence. Many computational tools implementing the ACMG guidelines have been developed (4–7). However, the degrees of subjectivity and uncertainty allowed in the guidelines have been found to lead to inconsistent and partly incompatible results (8).

Many studies and also clinical practice use gene panels of known predisposing genes to circumvent the challenges in variant interpretation. This approach does not allow identification of novel predisposing genes and restricts the detection of variants of unknown significance (VUS). Furthermore, the lack of well-matched population controls complicates high-quality evaluation of rare germline variants. This is exemplified when analyzing findings in populations underrepresented in genetic variant repositories such as gnomAD (*i.e.*, non-Europeans).

Despite successes in revealing the role of somatic genetic changes, hereditary predisposition in hematological malignancies has been acknowledged only recently. Reports typically focus on just one single hematological disease/malignancy or a gene (9). However, identification of pathogenic germline variants is crucial in hematology, as if timely recognized, they can direct therapeutic decisions, such as conditioning modalities or donor selection in allogeneic hematopoietic stem cell transplantation (HSCT) (10,11).

The Finnish population has been isolated, and has undergone genetic bottlenecks which left the population with a higher number of deleterious low-frequency variants compared to larger, mixed populations (12). This enrichment of variants allows powerful detection of genotype-phenotype associations of rare deleterious variants. In this study, we report a comprehensive pathogenicity evaluation of germline variants discovered in exome sequence data of 511 Finnish adult hematological patients (**Fig. 1**). To this aim, we developed PaVaDi, an automated pipeline implementing ACMG guidelines for variant pathogenicity classification. In gene panel level analysis we focused on genes associated with hematological diseases (HemMut) or DNA repair (DDR) and identified multiple P and LP variants. In an exome-wide burden analysis we identify three novel hematological malignancy predisposing gene candidates (*CUX2*, *RNPC3*, and *MFSD2A*) and affirm the role of *POT1*. We also demonstrate evidence for pathogenicity of certain VUSes in *MPO.* Finally, we solidify the significance of germline *CHEK2* variants doubling the risk for hematological malignancies.

**Figure 1.**
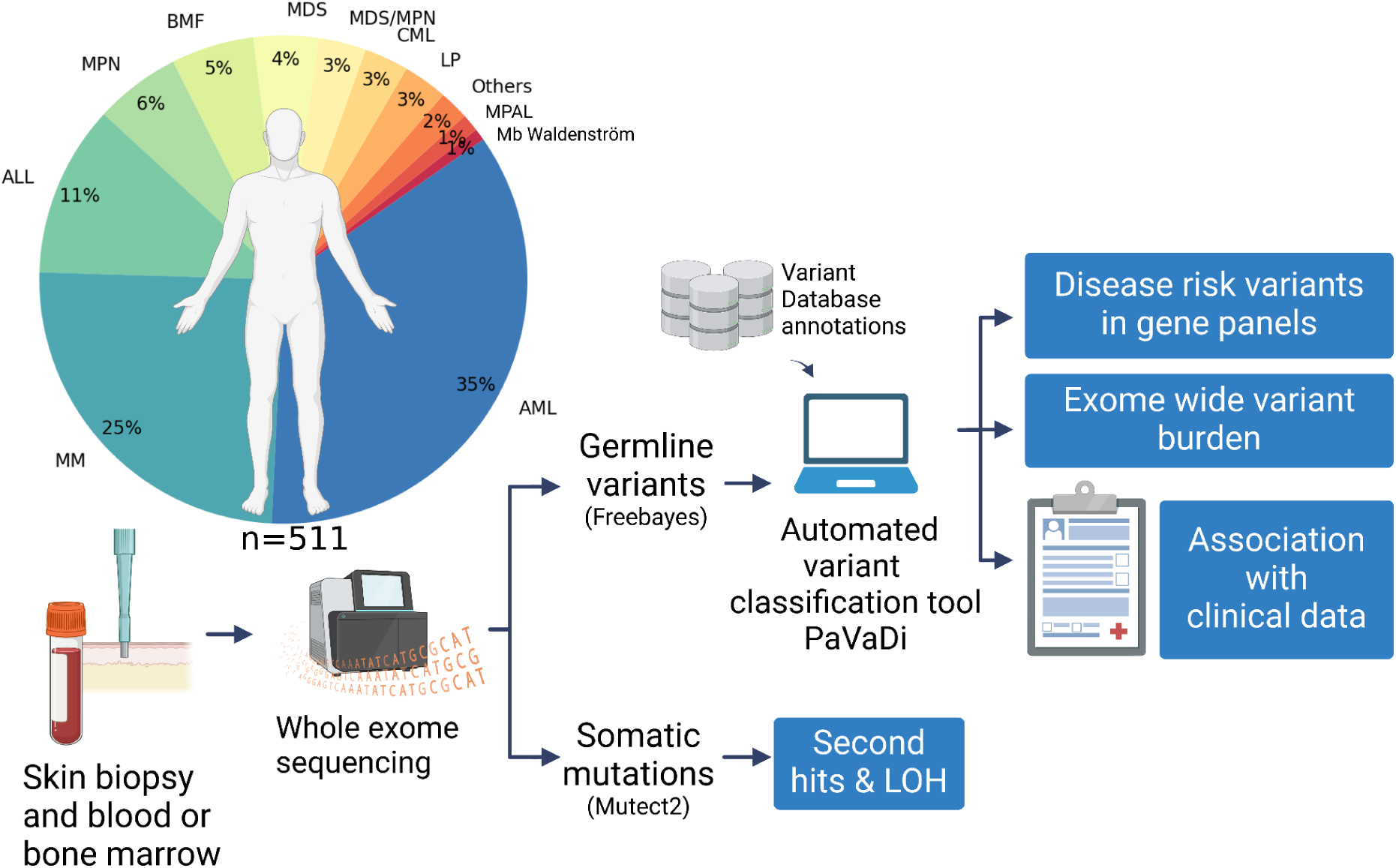
Overview of the sample set, study design and analysis workflow. AML, acute myeloid leukemia; ALL, acute lymphoid leukemia; BM, bone marrow; BMF, bone marrow failure; CML, chronic myeloid leukemia; MDS, myelodysplastic syndrome; MM, multiple myeloma; MPAL, mixed phenotype acute leukemia; MPN, myeloproliferative neoplasm; LOH, loss-of-heterozygosity; LP, chronic lymphoproliferative disease; Mb Waldenström, Waldenström’s macroglobulinemia.

## Methods

### Patients, samples, and exome sequencing

The study set consisted of 511 adult patients diagnosed with various hematological diseases, mostly malignancies in Finland. Patient characteristics were retrieved from hospital records and the Finnish Hematological Registry, and are summarized in Table 1 and in Supplementary Table 15. Samples were collected between 2011 and 2022 at the Department of Hematology of Helsinki University Comprehensive Cancer Center in (HUH CCC) and the Finnish Hematology Registry and Clinical Biobank (FHRB Biobank). Whole exome sequencing (WES) data was available for all 511 patients. Germline sequencing was performed on skin biopsy samples except for one remission on blood sample as described previously (13) with Nimblegen SeqCap EZ exome v2.0 (Roche, Basel, Switzerland), The SeqCap EZ MedExome (Roche, Basel, Switzerland), Twist Core Exome (Twist Bioscience, SF, USA) or Agilent Clinical Research Exome (Agilent, Santa Clara, CA, USA). In addition to the 511 germline WES, tumor WES was available for 460 patients (88.8%). Tumor samples were obtained from bone marrow (N=297), peripheral blood (N=31), CD138+ cells (N=121), CD3+; CD34+; CD4+/CD8+; CD8+ (N=1 in each), cerebrospinal fluid (N=1) or unknown primary site (N=6) at diagnosis, remission or relapse stage of the disease. Tumor WES had been performed as described previously (14). Both normal and tumor sequencing was performed at the Institute for Molecular Medicine Finland (FIMM) Genomics unit supported by HiLIFE and Biocenter Finland or at the Biomedicum Functional Genomics Unit (FuGU) at the Helsinki Institute of Life Science and Biocenter Finland at the University of Helsinki.

**Table 1.**
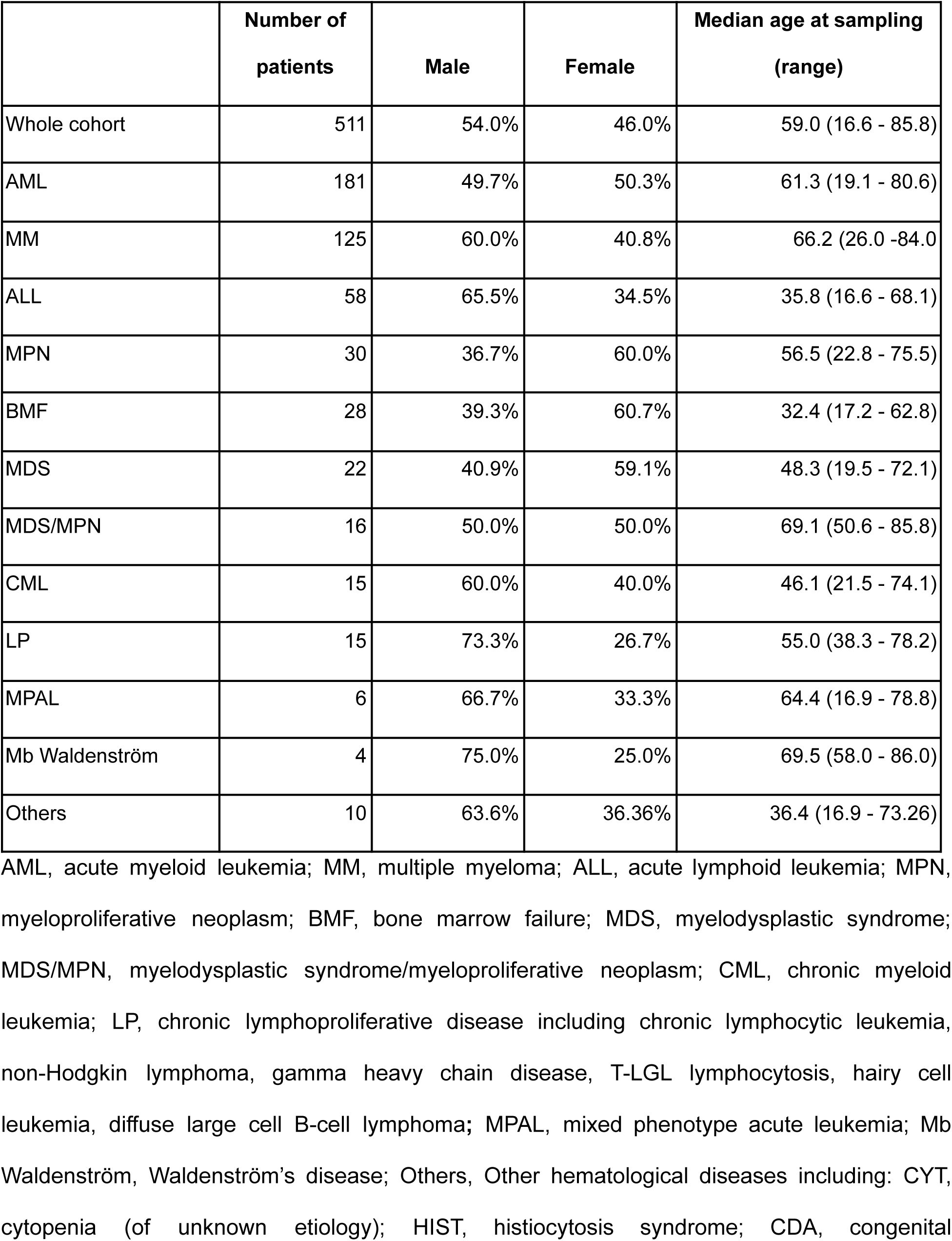
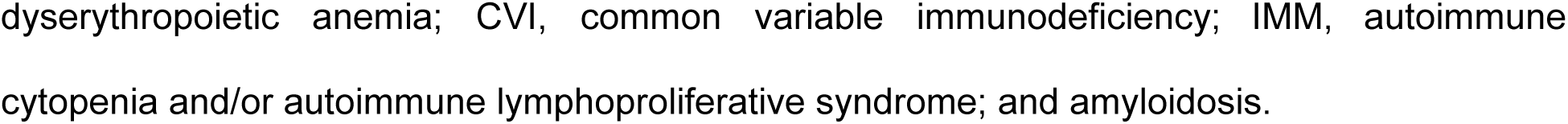
Patient characteristics.

|  | **Number of patients** | **Male** | **Female** | **Median age at sampling (range)** |
| --- | --- | --- | --- | --- |
| Whole cohort | 511 | 54.0% | 46.0% | 59.0 (16.6 - 85.8) |
| AML | 181 | 49.7% | 50.3% | 61.3 (19.1 - 80.6) |
| MM | 125 | 60.0% | 40.8% | 66.2 (26.0 - 84.0) |
| ALL | 58 | 65.5% | 34.5% | 35.8 (16.6 - 68.1) |
| MPN | 30 | 36.7% | 60.0% | 56.5 (22.8 - 75.5) |
| BMF | 28 | 39.3% | 60.7% | 32.4 (17.2 - 62.8) |
| MDS | 22 | 40.9% | 59.1% | 48.3 (19.5 - 72.1) |
| MDS/MPN | 16 | 50.0% | 50.0% | 69.1 (50.6 - 85.8) |
| CML | 15 | 60.0% | 40.0% | 46.1 (21.5 - 74.1) |
| LP | 15 | 73.3% | 26.7% | 55.0 (38.3 - 78.2) |
| MPAL | 6 | 66.7% | 33.3% | 64.4 (16.9 - 78.8) |
| Mb Waldenström | 4 | 75.0% | 25.0% | 69.5 (58.0 - 86.0) |
| Others | 10 | 63.6% | 36.36% | 36.4 (16.9 - 73.26) |

AML, acute myeloid leukemia; MM, multiple myeloma; ALL, acute lymphoid leukemia; MPN, myeloproliferative neoplasm; BMF, bone marrow failure; MDS, myelodysplastic syndrome; MDS/MPN, myelodysplastic syndrome/myeloproliferative neoplasm; CML, chronic myeloid leukemia; LP, chronic lymphoproliferative disease including chronic lymphocytic leukemia, non-Hodgkin lymphoma, gamma heavy chain disease, T-LGL lymphocytosis, hairy cell leukemia, diffuse large cell B-cell lymphoma; MPAL, mixed phenotype acute leukemia; Mb Waldenström, Waldenström's disease; Others, Other hematological diseases including: CYT, cytopenia (of unknown etiology); HIST, histiocytosis syndrome; CDA, congenital
dyseerythropoietic anemia; CVI, common variable immunodeficiency; IMM, autoimmune cytopenia and/or autoimmune lymphoproliferative syndrome; and amyloidosis.

### Population controls

GnomAD (v.2.1.1) global (n=125,748) and non-cancer Finns (n=10,816) (15) were used as population controls in variant level Fisher’s exact tests and gene level burden tests.

### Germline variant calling

To detect germline single-nucleotide variants (SNVs), multiple-nucleotide variants (MNVs) and short insertions or deletions (indels), variant calling was performed on normal tissue WES data using Freebayes (v.1.3.2) (16) in population sample calling mode with parameters -min-repeat-entropy 1, report-genotype-likelihood-max, -alternate-fraction 0.2, and -no-partial-observations. Raw variant calls were filtered to include only good quality variants in the set (filters for quality QUAL>20, QUAL/AO>2; strand bias artifacts SAF>1,SAR>1; read position artifacts RPR>1,RPL>1). Variants were normalized using bcftools (version 1.12) (17) and variants with multiple alleles were decomposed using vt (version 0.57) (18). Variants were further filtered on a sample level by including only calls with sample genotype quality (GQ) ≥ 20, coverage > 7 and allelic fraction (AF) ≥ 0.3.

### Somatic mutation calling

Somatic mutations were detected using Mutect2 (included in GATK (v4.2.0) (19)) in tumor-normal mode. A panel of normals was constructed for each sequencing kit used and gnomAD v.2.1.1 (15) was used as a germline variant resource. Somatic calls were filtered with GATK FilterMutectCalls. Only mutations with >4 supporting reads were included in the analysis.

### Evaluation of genetic ancestry

Genetic ancestry of the patients were evaluated by computing the first four principal components (PCs) of genotypes. Principal component analysis (PCA) was performed using the prcomp function in R (version 4.3.2). Prior to PCA, the variant data were pruned for linkage disequilibrium (LD) using PLINK software (version 2.0). LD pruning was performed with a window size of 50 single nucleotide polymorphisms (SNPs), a step of five SNPs and an r² threshold of 0.2. PCs were visualized to identify clusters corresponding to different ancestral populations. Eight patients were found to have non-Finnish ancestry and were removed from the exome-wide variant burden analysis.

### Gene panels

To examine genes previously associated with hematological disorders and DNA repair, we composed two panels of genes. The first panel consisted of 297 genes associated with inherited hematological disorders (HemMut; a panel developed for diagnostics at HUSLAB Laboratory of Genetics, HUS Diagnostic Center, Helsinki University Hospital) (Supplementary Table 1). The second panel consisted of 276 DNA damage repair (DDR) genes (Supplementary Table 2) (20).

### Inheritance patterns

Inheritance patterns were retrieved from Clinical Genomic Database (version December 2023) except for *SBDS, MYSM1* and *MPO* for which we annotated the inheritance patterns as AR, AR and AD/AR respectively.

### PaVaDi, a variant pathogenicity classification pipeline

The ACMG has published guidelines including 28 criteria for the clinical interpretation of germline variants with respect to human diseases (3). Following a rule-based strategy outlined in the guidelines, variants can be classified as pathogenic (P), likely pathogenic (LP), benign (B), likely benign (LB) or as variants of uncertain significance (VUS). There are different levels of pathogenicity supporting criteria: very strong (PVS1), strong (PS1-PS4), moderate (PM1-PM6) and supporting (PP1-PP5). Criteria supporting benign classification are stand-alone (BA1), strong (BS1-BS4) and supporting (BP1-BP7). If no criteria is met, or B/LB and P/LP evidence is contradictory, the variant is classified as VUS.

We developed an automated pipeline PAthogenic VAriant DIscovery (PaVaDi) that implements 17 of the criteria (PVS1, PS1, PM1, PM2, PM4, PM5, PP2, PP3, PP5, BA1, BS1, BS2, BP1, BP3, BP4, BP6, BP7) in addition to three strengthened criteria PM2_s, PP5_s, BP6_s that could be automatized with the data available to us (Supplementary Table 3; Supplementary Text). The pipeline consists of an annotation module scoring each variant with a set of rules, each 1 or 0, based on the available evidence, and a classification module assigning a pathogenicity level (B, LB, VUS, LP or P) based on the scored rules (Supplementary Figure S2).

We integrated multiple tools and databases that can be used to assess the significance of genetic variants to disease into the pipeline (Supplementary Table 4). They provide evidence on variant pathogenicity for the criteria, upon which the pipeline is assigning the variants a pathogenicity level (B, LB, VUS, LP or P). We used variant and gene lists from the InterVar package for evidence in the scoring of criteria (21).

### Exome-wide variant burden analysis

We studied the enrichment of rare variants in the patient set compared to gnomAD Finns without a cancer diagnosis. We performed a burden test with the Total Frequency Test (TFT) (22), collating rare P+LP, B+LB, and VUS variants at the gene level in each pathogenicity class separately. To alleviate potential detection bias due to differences in sequencing technologies and data analysis methods between the patient set and the population controls, we included only non-synonymous variants which were present in both the patient set and controls, and altogether had at least three variant alleles in the patient set and controls combined. Genomic inflation in the data set was corrected by dividing χ² statistics with λ using P/LP, B/LB and VUS burden per gene, where λ is the observed median χ² statistic divided by the expected median χ² statistic (Supplementary Figure S3-5). Benjamini-Hochberg procedure was used for multiple testing correction with a false discovery rate (FDR) threshold at 10%. All variants with FDR less than 10% were visually confirmed in sequencing data with BasePlayer (23).

We studied the enrichment of variants across different pathogenicity classes by comparing the minor allele frequency (MAF) in the study set to the MAF in gnomAD non-cancer Finns or gnomAD non-cancer total, selecting the higher MAF of these two as the frequency in controls. Only variants with more than zero alleles in both cases and control were included to alleviate potential detection bias. Fisher’s exact test (two-sided) was used to determine how likely it is that the variant enriched in cases (over twofold enrichment in cases compared to control) is P/LP.

### Linear regression analysis

We employed ordinary linear regression models, with intercept, using Python package statsmodels to predict 1) AlphaMissense variant pathogenicity score and 2) variant enrichment in the patient set (log_2_(MAF(case) / MAF(control)) with ACMG rules as predictors.

### *In silico* protein stability prediction

We used DynaMut and mCSM (24) to assess the impact of missense mutations on protein stability. Protein stability alterations were determined as differences in free energy (ddG). DdG ≥ 0 was defined as stabilizing and ddG < 0 as destabilizing. In addition, we used AlphaMissense (25) to assess whether pathogenicity predictions support the protein stability analysis results. Protein structures and mutations were visualized with PyMol (The PyMOL Molecular Graphics System, Version 2.5.0 Schrödinger, LLC).

### *In silico* splicing prediction

We used SpliceAI (26) to predict the probability that *CUX2* splice variant c.1259-2A>G disrupts splicing. SpliceAI outputs delta-scores (values between 0 and 1) for the probability. Cutoff 0.8 is considered high precision.

### Loss of heterozygosity and biallelic events

We estimated loss-of-heterozygosity (LOH) at variant loci by testing whether the variant allelic fraction (VAF) was significantly higher in tumor than in normal using Fisher’s exact test for the ratio VAF^tumor^ / VAF^germline^ for each variant. The resulting *p*-values were adjusted with the Benjamini-Hochberg procedure, and events with FDR<1% were considered LOH.

For biallelic events, we examined genes carrying both a P/LP germline variant and either a missense or a truncating somatic mutation.

### Post-translational modification analysis

We mapped germline missense variants to post-translational modification sites (PTM) using ActiveDriverDB (27). PTM site substitutions were divided into four categories: direct substitutions replaced the amino acid affected by post-translational modification, proximal substitutions replaced amino acids within ±2 and distal substitutions replaced amino acids within ±7 amino acids of the PTM site, and network-rewiring substitutions replaced amino acids near PTM site in kinase binding motif.

## Results

### Evaluation of germline variant pathogenicity in 511 patients with a hematological disease

In order to assess the pathogenicity of a large number of germline variants in whole exome data, we developed PaVaDi, an automated pipeline integrating evidence available in databases such as ClinVar, and allele frequencies (AF) in population databases to automatically assign each variant with one of five pathogenicity classes (B, LB, VUS, LP, P) based on 17 of the rules described in the ACMG guidelines (3) (Methods; Supplementary Table 3; Supplementary Figure S2). PaVaDi is freely available at https://github.com/jezkoski/PaVaDi.

We discovered 156,245 germline variants (**Fig. 2 a-c**) in our study set of 511 patients with hematological disease diagnosed in Finland (Methods). We assigned each variant a pathogenicity class (“ACMG verdict”) with PaVaDi, resulting in 54,381 benign (B, 34.8%), 54,920 likely benign (LB, 35.1%), 45,289 (VUS, 29.0%), 1,224 likely pathogenic (LP, 0.8%) and 431 pathogenic variants (P, 0.3%) (**Fig. 2 a**). As expected, B/LB variants showed higher allele frequencies than P/LP variants in both cases and controls (**Fig. 2 b-c**), and variants rare in population showing higher frequencies in cases (**Fig. 2 d**). The latter skew was partly (34%) attributable to case-control imbalance (Supplementary Text, Supplementary Figure S1), with the remainder likely stemming from differences in variant analysis workflows and presence of true pathogenic variants. Moreover, the variants which were more than two-fold enriched in our study set were more frequently P/LP/VUS variants than in the Finnish population (5.4-fold, *p*<2.2×10^−16^, 95% CI 5.1–5.8; **Fig. 2 f**; Supplementary Text). Multiple ACMG rules were found to correlate with enrichment (**Fig. 2 h**; Methods). As expected, these included rules for higher (PM2) or lower frequency in controls (BS1, BA1), but also rules denoting a known mechanism or mutation hotspots (PVS1, PM1), and evidence of (non-)pathogenicity (BP6, PP3). The P/LP/VUS variants were also enriched in the Finnish population over the global population (5.4-fold, p<2.2×10^−16^, 95% confidence interval 5.2–5.6; **Fig. 2 e**; Supplementary Text), compatible with previous studies (12).

**Figure 2.**
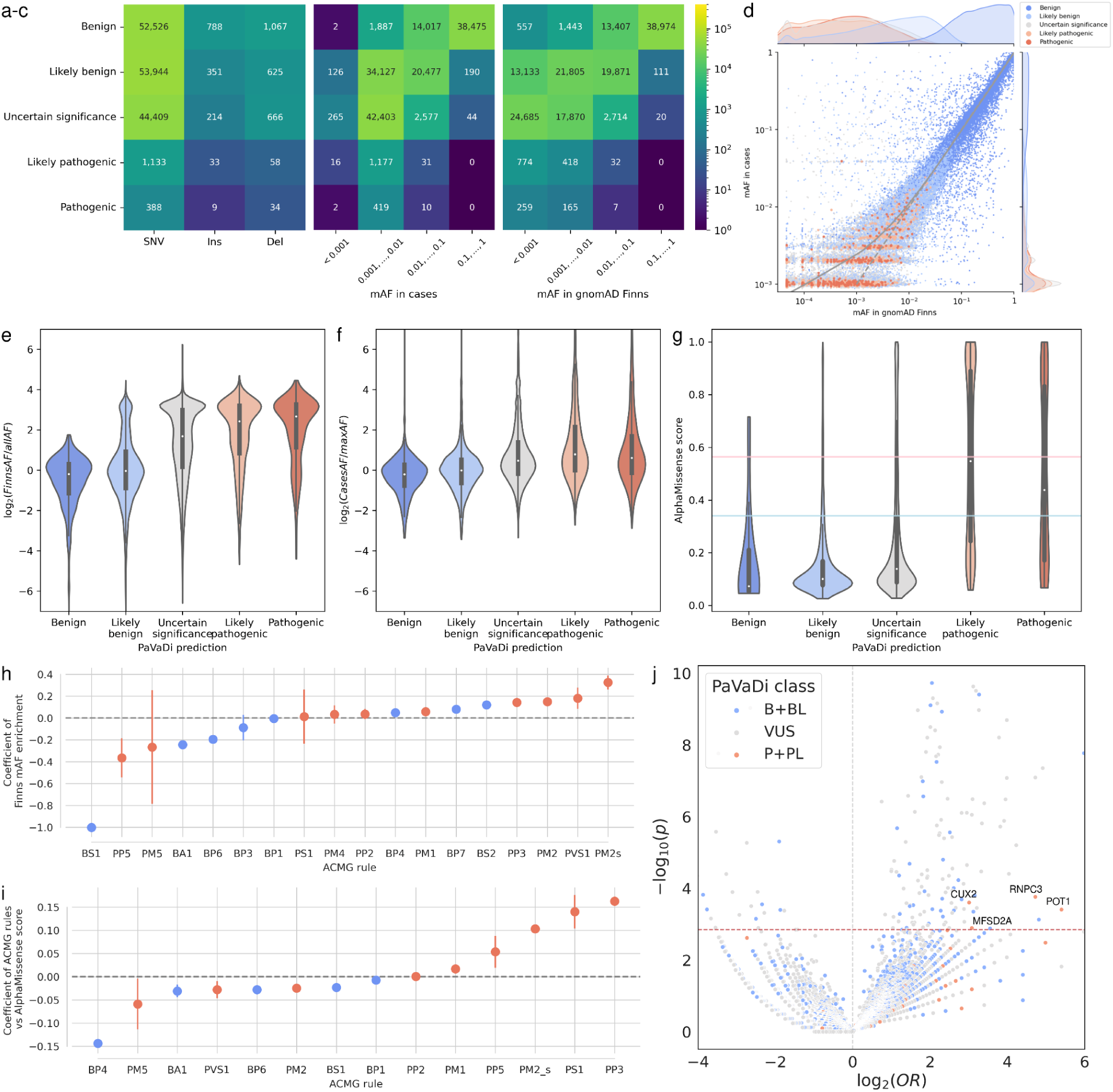
Germline variants (*n*=156,245) discovered in the hematological patient cohort. Distribution of germline variants by variant pathogenicity class assigned by PaVaDi, and **a**) variant type, **b**) MAF in cases and **c**) MAF in controls. **d)** A scatterplot of MAF in controls (X-axis) and cases (Y-axis) stratified by pathogenicity class. Solid line shows a local regression (LOESS) fit. **e)** Variant allele frequency enrichment stratified by pathogenicity class. Ratio of MAFs in gnomAD Finns (FinnsAF) vs gnomAD total (*n*=92,855). **f**) Ratio of MAFs in the hematological patient cohort (CaseAF) vs maximum MAF in gnomAD Finns and gnomAD total (*n*=101,225) stratified by pathogenicity class. **g)** AlphaMissense pathogenicity predictions stratified by pathogenicity class. Light blue line denotes the threshold for benign and pink for pathogenic prediction. **h)** Coefficients from a linear regression model explaining variant enrichment in cases with ACMG rules. Pathogenic and benign rules denoted by red and blue colors, respectively. **i)** Coefficients from a linear regression model of P/LP variants explaining AlphaMissense score with ACMG rules. **j)** Volcano plot of exome-wide variant burden analysis showing variant enrichment (log_2_(OR); X-axis) and significance (−-log_10_(p); Y-axis) stratified by pathogenicity class. Genes with significant P/LP variant burden at (FDR<11%) are labeled.

We then investigated the correlation of our pathogenicity classification with protein folding based predictions of functional effects for missense variants by AlphaMissense (25) in our study set. For P and LP variants, we observed a bimodal distribution of AlphaMissense scores, with 70% of P and 48% of LP variants being classified as pathogenic, whereas a majority of B/LB/VUS variants were classified as benign (**Fig. 2 g**). High AlphaMissense scores associated with pathogenic ACMG rules PP3, PS1, PM2s, PP5 and PM1, but interestingly not with PM5 denoting a novel missense at an amino acid residue where another missense has been previously determined pathogenic (**Fig. 2 i**; Methods).

### Gene panels highlight *CHEK2* in hematological malignancy predisposition

We performed an inspection of P/LP variants in genes that have previously been associated with predisposition to hematological disorders (HemMut panel, 297 genes), or DNA repair and replication (276 genes; Methods).

Examining the genes in the HemMut panel (Supplementary Table 1, Methods), we found 274 P/LP variants out of a total of 2,392 variants. In total, 115/511 (22.5%) individuals carried P/LP variants in AD, AD/AR, biallelic AR, or XL (males) genes (**Fig. 3**; Supplementary Table 5-7). Five individuals carried biallelic mutations in recessively inherited *ERCC6L2* (n=4, 0.7%) and *CTC1* (n=1, 0.2%) genes. The most frequently mutated genes were *CHEK2* (n=19 patients, 3.7% of all patients), *FANCM* (n=14, 2.7%), *CTC1* (n=13, 2.5%), *TRNT1* (n=12, 2.3%), *ANO6* (n=9, 1.8%), *GP6* (*n*=9, 1.8%), and *VWF* (*n=*9, 1.8%). Seven female individuals were identified with a hemizygous variant in three X-linked genes *ALAS2* (n=3), *ATRX* (n=1), and *F8* (n=3). Two of the *ALAS2* variant carriers had previously been diagnosed with congenital dyserythropoietic anemia. Moreover, we observed six P/LP variants enriched in the Finnish population (2-12 fold) to be more prevalent among patients than expected (Table 2).

**Figure 3.**
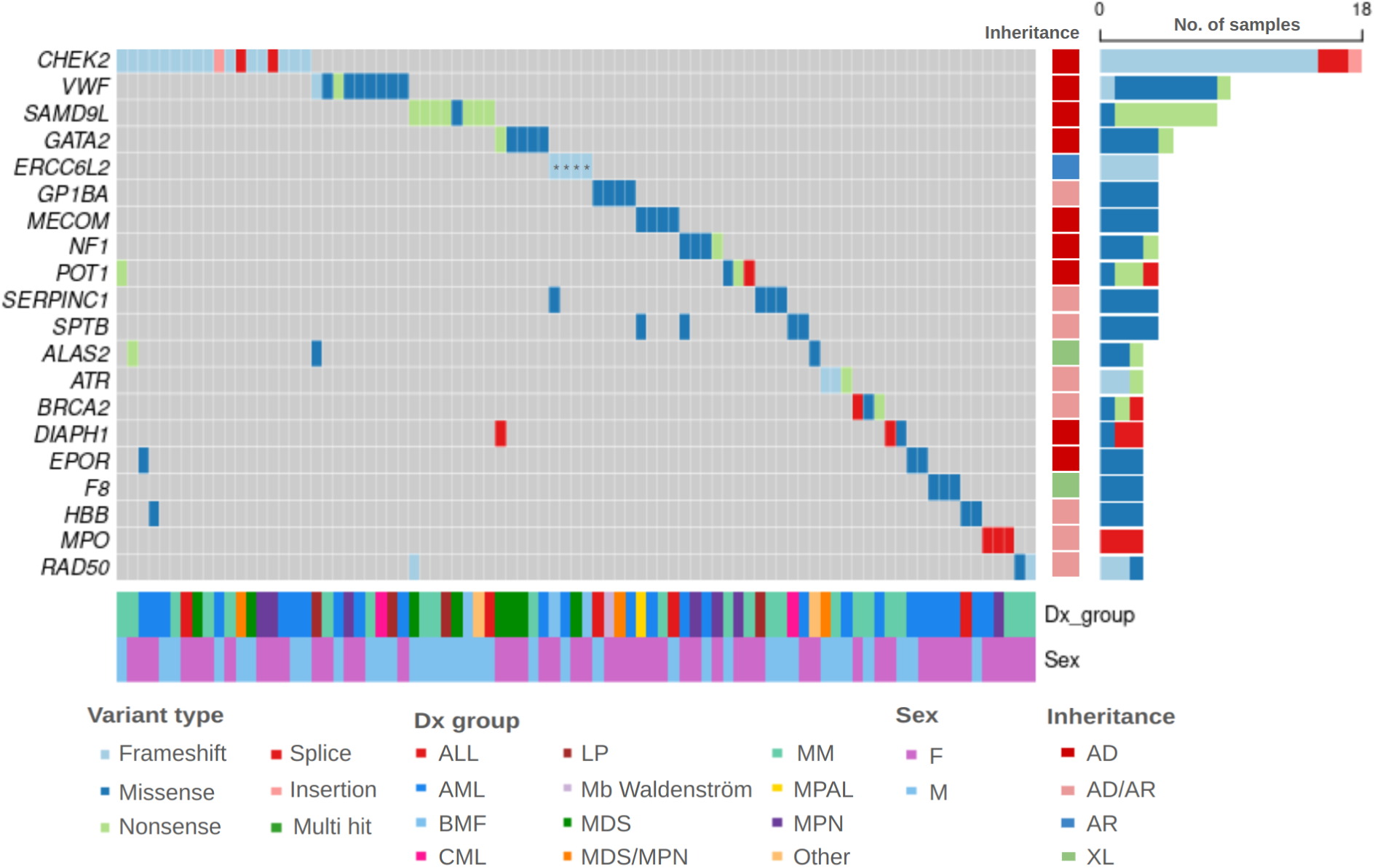
Individuals with the most frequent heterozygous AD and XL, and homozygous/compound heterozygous AR P/LP variants in 20 genes of the HemMut panel (Y-axis) for 98 carriers (X-axis). Homozygous AR gene variants marked with *. Columns on the right indicate the mode of inheritance and the total number of variants colored by variant type. Rows on the bottom indicate diagnosis group and sex for each patient.

**Table 2.**
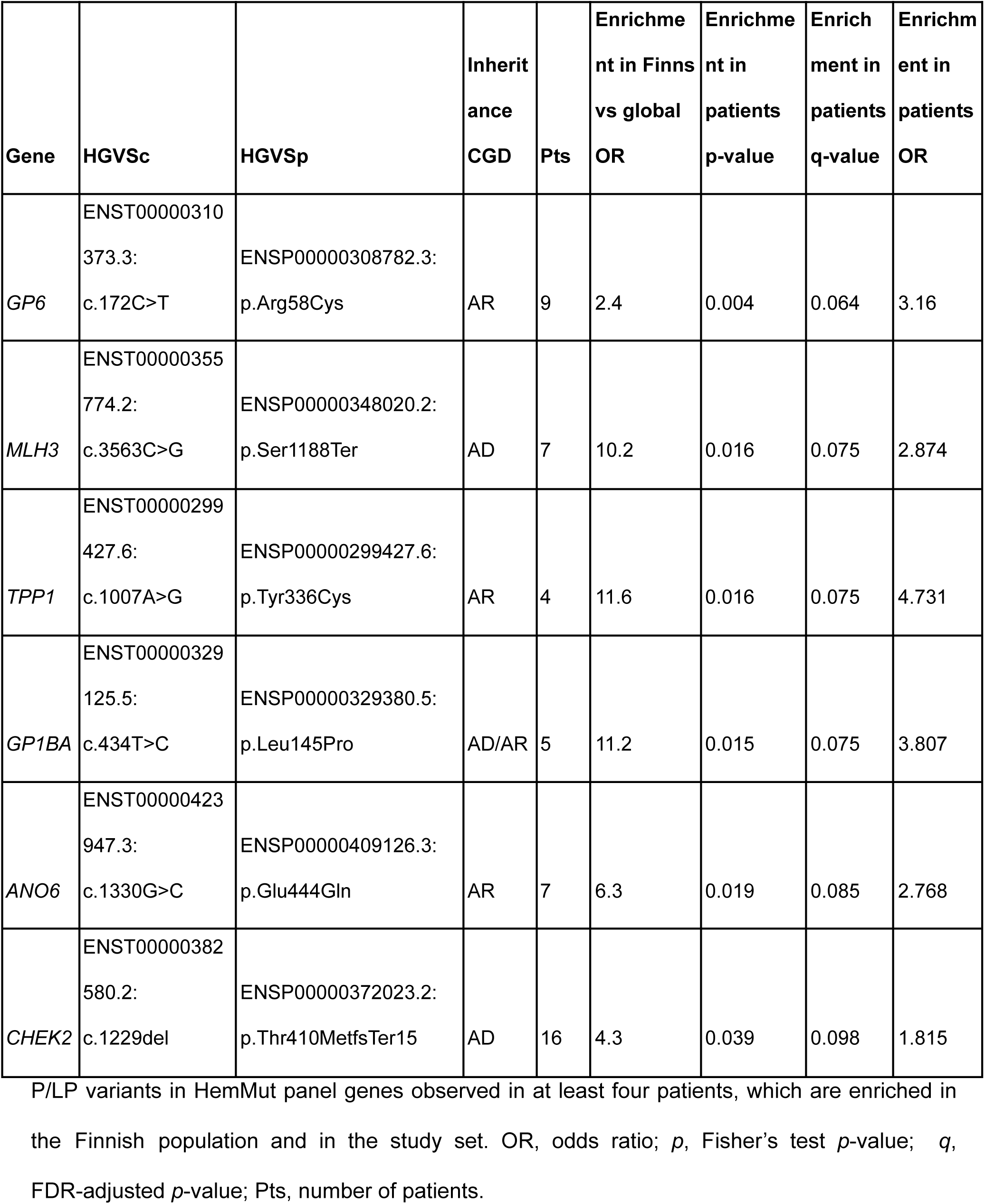
Finnish enriched P/LP variants in the HemMut panel.

Inspecting the DDR genes (DDR panel, Methods), we found 12.3% (63/511) of patients with at least one rare P or LP heterozygous AD, AD/AR, or biallelic autosomal AR variant in 21 genes (**Fig. 4**; Supplementary Table 7-8; Supplementary Table 5). *CHEK2* variants were the most frequent also in this panel. We then studied heterozygous variants in AR genes of the DDR panel as some of them have been shown to confer risk even at heterozygous state (12,28) (Supplementary Table 7). The most frequently mutated genes were *FANCM* (*n*=14), *ERCC8* (*n*=10), *MSH5* (*n*=6), *MUTYH* (*n*=6) and *RNF168* (*n*=5). Analysis of somatic secondary events revealed four genes (*FANCM*, *ATM*, *LPIN2* and *DDX41*) where a P/LP germline variant was coupled with a missense or truncating somatic mutation in the same gene (Supplementary Text; Supplementary Table 14).

**Figure 4.**
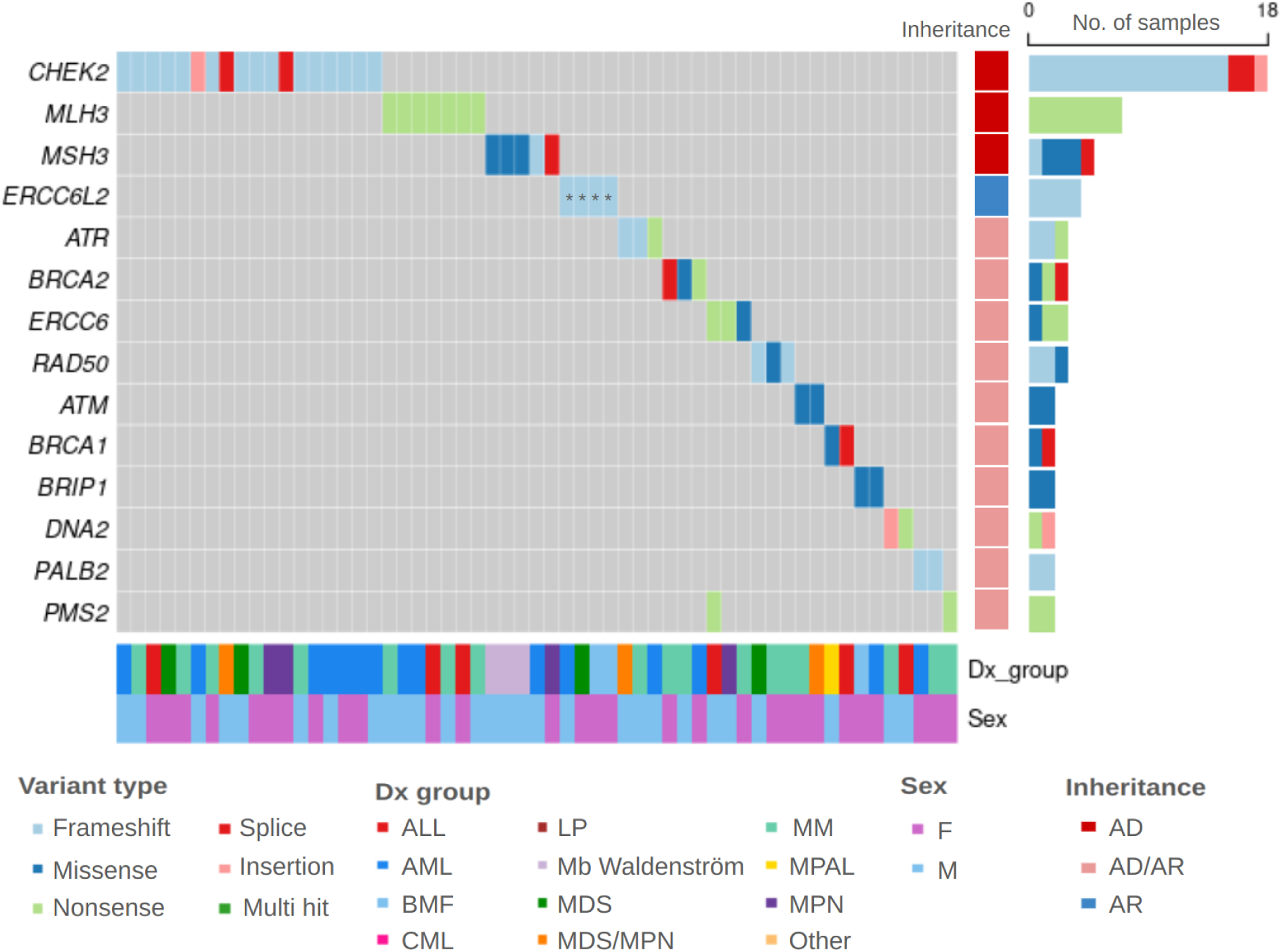
Heterozygous AD and homozygous AR P/LP variants in 14 genes of the DDR panels (Y-axis) for 57 carriers (X-axis). Only genes with two or more variants are shown. Homozygous AR gene variants marked with *. Columns on the right indicate the mode of inheritance and the total number of variants colored by variant type. Rows on the bottom indicate diagnosis group and sex for each patient.

### Germline *CHEK2* variants are enriched in both myeloid and lymphoid hematological malignancies

We then examined *CHEK2* variants in more detail, and identified 19 P/LP variants (c.1497dup, c.1229del, c.319+2T>A) and four VUSes (Asp481Tyr, Glu282Lys). The known founder variant Ile200Thr was classified as LB in our pipeline due to the high MAF among Finns (Supplementary Table 10). However, we included it in our analyses given the evidence of a predisposing role in *e.g.* MPNs (29). In total, 14% (71/511) of patients were carriers of one of six unique *CHEK2* variants (OR=1.78, CI 95% 1.35-2.30, *p*=4.5×10^−5^) (**Fig. 5**; Supplementary Table 11). The most prevalent variant Ile200Thr was enriched at the whole cohort level (*n*=49 carriers, 10%, OR=2.04, CI 95% 1.47–2.78, *p*=2.0×10^−5^) and, more specifically, in both ALL (19%, 11/58, OR=4.5, CI 95% 2.09–8.86, *p*=0.00012) and AML patients (9%, 16/180, OR=1.87, CI 95% 1.04–3.17, *p*=0.024). The second most frequent variant was c.1229del, which was found in 3% of patients (16/511, OR=1.81, CI 95% 1.01–3.05, *p*=0.039). We also identified one myeloma patient (JK202) with LOH (FDR=0.002) at this locus. One patient (JK184) carried two variants in *CHEK2*: a pathogenic frameshift variant c.1497dup and a VUS missense variant Glu282Lys.

**Figure 5.**
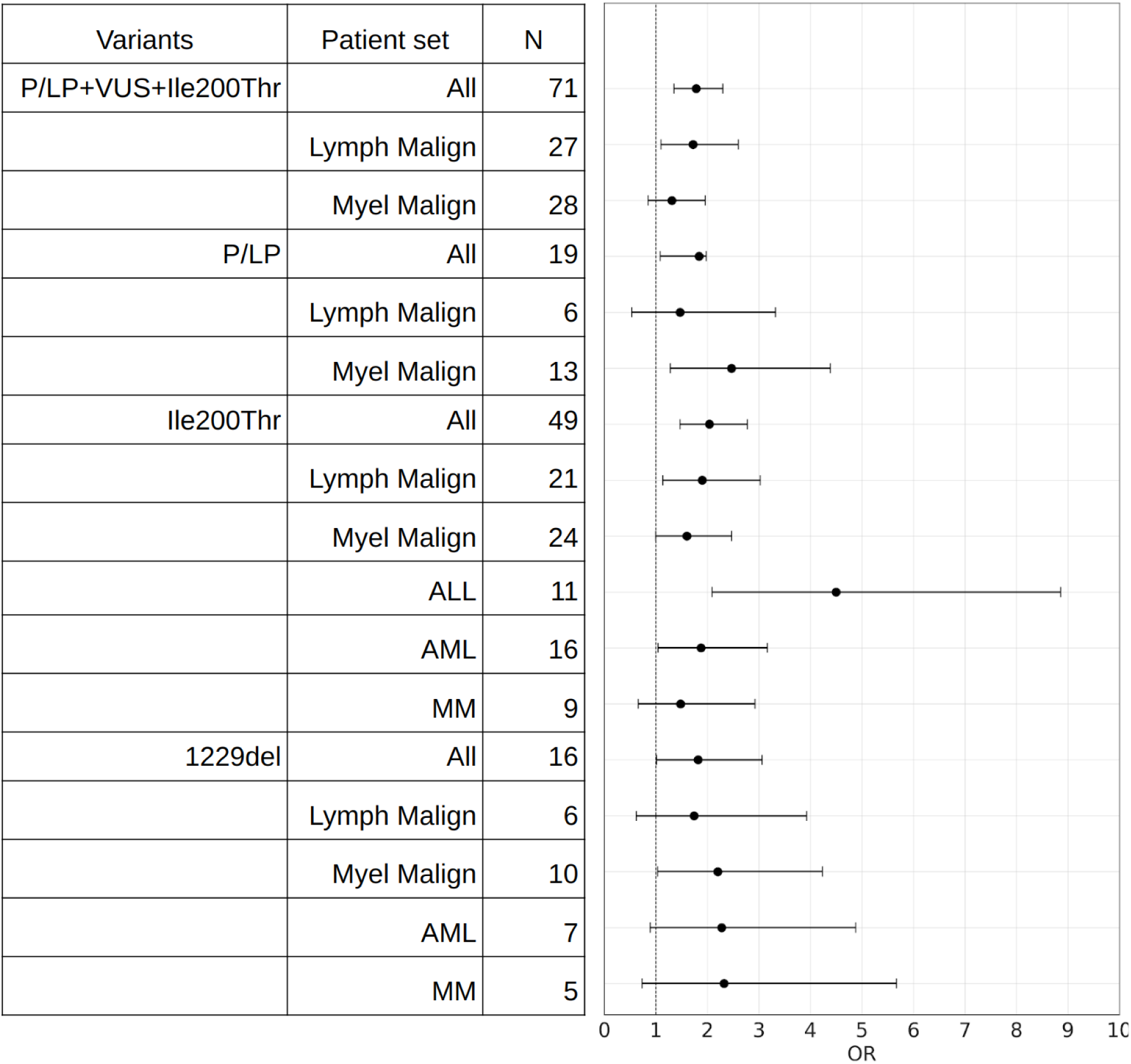
A forest plot showing ORs and 95% confidence intervals of P/LP (c.1497dup, c.1229del, c.319+2T>A), VUS (Asp481Tyr, Glu282Lys) and Ile200Thr *CHEK2* variants in the whole patient cohort and subgroups compared to population control gnomAD Finns. Only groups with five or more variants are shown. Lymph Malign patient set includes ALL, MM and LP. Myel Malign patient set includes AML, MPN, MDS, MDS/MPN and CML.

### Exome-wide germline variant burden analysis suggests novel predisposing genes *CUX2, RNPC3,* and *MFSD2A*

We next asked whether the study set harbored previously uncharacterized risk alleles missed in the panel analyses. To this end, we performed an exome-wide burden test to detect genes with more P/LP germline variants than expected in the patient study set with 503 patients of Finnish ancestry (Methods). Four genes, *CUX2*, *RNPC3*, *MFSD2A* and *POT1,* harbored more P/LP variants in the patient set compared to population controls (OR>2.7, gnomAD Finns non-cancer, FDR<10%; **Fig. 2 j**; Table 3; Supplementary Table 9; Supplementary Figure S3-4). The most frequent variant observed in *CUX2* was splice variant c.1259-2A>G (*n=*6). One of the patients was homozygous for this variant. Four out of seven *CUX2* variant carriers with known family history had first-degree relatives with cancer (Supplementary Table 9). SpliceAI predicted the variant to be splicing disturbing (probability 0.97; Methods).

**Table 3.** P/LP variant burden test.

| Gene | Cases | Controls | OR | 95%<br>confidence<br>interval | P-value | $\lambda$ adjusted<br>p-value | λ-adjusted<br>q-value |
| --- | --- | --- | --- | --- | --- | --- | --- |
| *RNPC3* | 5 | 4 | 26.4 | 5.7–133.6 | $2.1 \times 10^{-5}$ | 0.0002 | 0.0153 |
| *CUX2* | 8 | 21 | 8.1 | 3.1–19.1 | $3.3 \times 10^{-5}$ | 0.0003 | 0.0218 |
| *POT1* | 4 | 2 | 42.3 | 6.0–466.4 | $5.9 \times 10^{-5}$ | 0.0004 | 0.0325 |
| *MFSD2A* | 6 | 15 | 8.5 | 2.7–23.2 | 0.00026 | 0.0013 | 0.0910 |

Results of a test for P/LP variant burden of $n=1,552$ genes in the patient study set. Results at $FDR < 10\%$ are shown. Cases, number of patients carrying a P/LP variant; Controls, number of variant carriers in gnomAD Finns non-cancer controls; $p$ , p-value for the burden test; $\lambda$ adjusted $p$ , p-value adjusted for inflation (Methods); $\lambda$ adjusted $q$ , p-value adjusted for inflation and multiple testing correction with Benjamini-Hochberg method.

Two other variants observed in the gene were Arg50Gln (*n=*1) and Asp307Tyr (*n=*1). We also identified five patients with three distinct *RNPC3* variants: Asp250Gly (n=3), Gln432Arg (n=1), and Glu229Val (n=1) and seven patients with three unique *MFSD2A* variants: Arg366Trp (*n*=3), Tyr444Cys (*n*=2), and Pro476Ala (*n*=1). We have previously reported the four *POT1* variants in association with myeloma predisposition (30).

### Novel germline variants in *MPO* are predicted to destabilize the protein

*MPO* has only recently been proposed as a predisposing gene to myeloid malignancies in heterozygous state (31)). We thus examined also VUSes in *MPO* aiming to identify novel potentially pathogenic variants. In our study set three patients carried a pathogenic splice variant (c.2031-2A>C; Supplementary Table 12) previously reported in MPO deficiency syndrome (32), and as a predisposing variant to myeloid malignancies (31). Furthermore, eight patients harbored heterozygous VUS variants. Four of the patients carried missense variants (Arg548Trp in two patients, Arg590Cys, Arg460Gln) previously reported as LP in (31,33) and four patients novel VUSes (Supplementary Table 12). Of these novel variants, Ile640Phe showed LOH (FDR=0.5%) while Asn716Ile is near a phosphorylation site (PTM analysis, Methods). Both of these and the other two VUSes (Thr417Ile and Trp255Arg) are located in conserved positions (phyloP100way_vertebrate>6 and >9, respectively).

In our patient set, the detected *MPO* variants were not limited to patients with myeloid malignancies, but occurred also in patients diagnosed with multiple myeloma, T-cell large granular lymphocytosis, and bone marrow failure (BMF) (Supplementary Table 12). All but one of the nine distinct *MPO* variants identified (P/LP *n*=3; VUS *n*=8) are located in the myeloperoxidase-like domain (**Fig. 6 e**).

**Figure 6.**
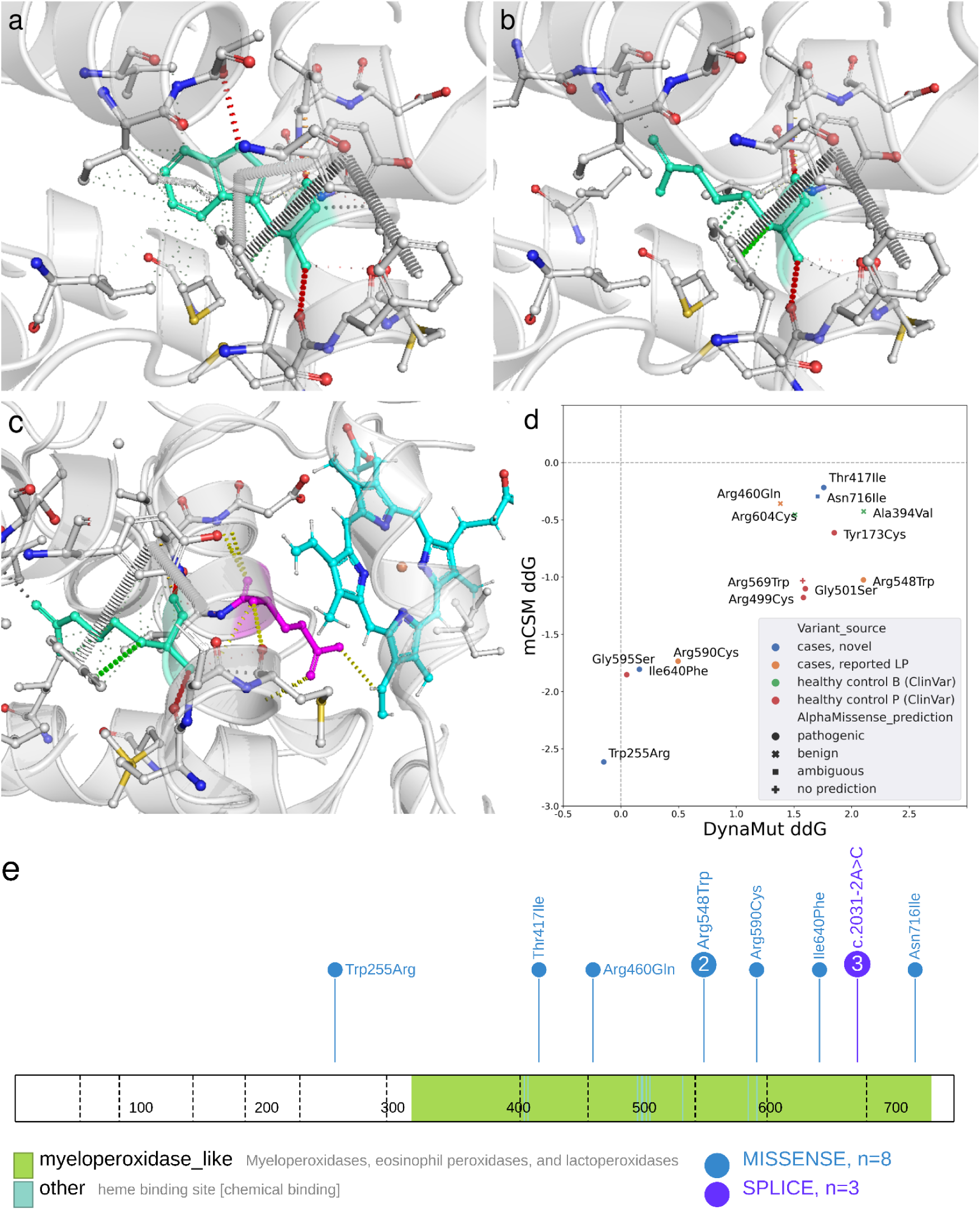
Protein stability analysis of *MPO* variants. Amino acid structures and bonds with neighboring residues of wild type Trp255 (**a**) and Trp255Arg (**b**) predicted with DynaMut (24). **c**) Trp255Arg (turquoise) and a catalytic residue Gln257 (pink) are located in the same alpha helix. **d**) Protein stability and pathogenicity predictions for *MPO* variants identified in our analysis (cases) as well as for population variants (gnomAD) that were classified as B/LB or P/LP in ClinVar. Negative and positive ddG predicted by DynaMut and mCSM for the missense mutation imply destabilizing and stabilizing effects, respectively, on the protein structure stability. *MPO* variants are grouped as follows: cases, novel=Novel variants in cases; cases, reported LP=Variants reported earlier to be LP; control B=gnomAD variants labeled as benign in ClinVar; control P=gnomAD variants labeled as pathogenic in ClinVar. AlphaMissense pathogenicity predictions are denoted by shape. Threshold (ddG<0) for destabilizing prediction for both DynaMut and mCSM in gray dotted line. **e**) Variants found in the patient cohort distributed across *MPO* protein.

We then evaluated the effect of these variants on MPO stability with DynaMut and mCSM (24). DynaMut and mCSM scores showed high correlation (*r*=0.86, *p*=9.48×10^−5^). mCSM predicted all variants to be destabilizing (Supplementary Table 13, **Fig. 6 d**). However, DynaMut predicted all variants except Trp255Arg to be stabilizing. DynaMut predicted Trp255Arg to lead to substantially fewer bonds to the nearby residues (**Fig. 6 a, b**), with a potential effect on the catalytic residue Gln257 in the same alpha helix (34) (**Fig. 6 c**). Finally, analysis with AlphaMissense (**Fig. 6 d**) supported the pathogenicity of the novel variants discovered here.

## Discussion

Founder populations such as the Finnish population offer unique advantages to discover pathogenic germline variants (12). Here, we performed a pan-hematological analysis to discover and characterize possible hereditary risk variants in known and unknown predisposition genes. The strength of this study is that our analyses are all based on matched population controls enabling accurate risk stratification.

We developed a computational pipeline PaVaDi that allowed us to classify all 156,245 germline variants discovered in the study cohort by pathogenicity, resulting in 1% of variants being classified either pathogenic or likely pathogenic. To extract the clinically important and validated germline variant incidence in hematological patients, we first focused on variants in genes with a known role in hematological diseases or DNA damage repair. We demonstrate that one fifth of the study patients carried a P/LP germline variant. This is in line with our expectations considering the spectrum of the genes and diagnoses of study patients analyzed.

Strikingly, in both panels analyzed *CHEK2* emerged as the gene with the most variants. Powered by the high frequency of *CHEK2* variants (n=71) among our study patients we were able, for the first time, to demonstrate a well-quantified enrichment of these variants in a pan-hematological setting. *CHEK2* variants conferred a two-fold risk for HMs as a whole which is compatible with previous findings examining myeloid cancers (29,35). In our study, the risk for myeloid malignancies was approximately two-fold. Intriguingly, *CHEK2* Ile200Thr associated with 4.5-fold risk for ALL. This exceeds the risk for solid cancers previously reported for this or any *CHEK2* germline variant (36,37).

As predisposition genes for ALL are much less studied and acknowledged in both scientific literature and clinical practice than myeloid malignancies, our discovery has implications to comprehensive ALL care. Our ALL study population consisted only of adult ALL patients further increasing the impact of the finding: 20-50% adult patients with ALL undergo allogeneic hematopoietic stem cell transplantation as routine care. Using sibling donors with a potential carriership of *CHEK2* variants endangers patients with a donor-derived hematological malignancy of any kind. Even though penetrance of the *CHEK2* Ile200Thr seems incomplete, the risk for secondary HM is evident as the disease has already once been realized. Importantly, NGS panels used in diagnostics for ALL (or myeloid malignancies) do not routinely include *CHEK2*. We suggest this practice to be modified in the future. In Finland, we also face additional challenges as Ile200Thr is exceptionally frequent among Finns (2.8%), yet still clinically unappreciated.

We also scrutinized the exome-wide pathogenic variant burden which highlighted three novel genes, *CUX2*, *RNPC3*, and *MFSD2A,* not previously associated with hematological diseases. Although *CUX2* variants have been reported in breast cancer (38), its potential role in hematological malignancies remains uncharacterized. *CUX1*, a paralog of *CUX2*, is typically impaired in myeloid malignancies and acts as a tumor suppressor (39). Further validation is needed in order to confirm the role of these candidate genes in HMs.

VUSes present an important challenge in clinical practice. In our study, 29% of variants remained VUS after PaVaDi analysis, each patient carrying an average of 116 VUSes. As a proof of concept to resolve such variants, we examined VUSes detected in *MPO*, which has been recently associated with HM predisposition (31). Multiple computational tools supported the pathogenicity of previously unreported variants Trp255Arg and Ile640Phe. However, there is still considerable discordance between different tools for assessing VUSes in general, posing a barrier to obtaining clinically useful information. The resolution of VUSes is paramount for clinical practice because it directly impacts patient care and clinical decision-making. Inability to find causative variants leaves patients and clinicians in a state of diagnostic uncertainty, limiting actionable interventions such as targeted therapies, surveillance strategies, or familial risk assessments. Furthermore, the accumulation of unresolved VUSes in genetic databases can complicate the interpretation of future variants for meaningful clinical insights.

Our findings highlight the advantages of conducting genetic studies within founder populations such as enhanced detection of pathogenic variants. However, the relevance and impact of these discoveries extend far beyond these populations, offering valuable insights into cancer predisposition on a global scale. By identifying pathogenic variants linked to hematological malignancies, we contribute to the growing body of evidence supporting the critical role of inherited genetic risk factors in the development of these diseases. These insights not only enhance our understanding of the genetic underpinnings of hematological malignancies but also have direct implications for clinical practice, including opportunities for early detection, targeted interventions, and improved patient care.

## Conclusions

We have identified pathogenic germline variants across different hematological diseases identifying novel predisposition candidate genes and variants, and underscoring the importance of *CHEK2* variants particularly in ALL.

## Data Availability

All unidentifiable data is available within the article and its supporting files. The individual-level sequencing data cannot be made publicly available because the research participant consent
does not include authorization to share identifiable data.

## List of abbreviations

ACMG: American College of Medical Genetics and Genomics
AML: Acute myeloid leukemia
ALL: acute lymphoid leukemia
B: Benign
BM: Bone marrow
BMF: Bone marrow failure
CDA: Congenital dyserythropoietic anemia
CML: Chronic myeloid leukemia
CVI: common variable immunodeficiency
DDR: DNA damage response and repair
FDR: False-discovery rate
HIST: Histiocytosis syndrome
HM: Hematological malignancy
IMM: Autoimmune cytopenia and/or autoimmune lymphoproliferative syndrome
HSCT: Hematopoietic stem cell transplantation
LB: Likely benign
LOH: Loss-of-heterozygosity
LP: Likely pathogenic
LP: Chronic lymphoproliferative disease
Mb Waldenström: Waldenström’s macroglobulinemia
MDS: Myelodysplastic syndrome
MM: Multiple myeloma
MPA: Mixed phenotype acute leukemia
MPN: Myeloproliferative neoplasm
OR: Odds ratio
P: Pathogenic
VUS: variant of unknown significance

## Declarations

### Ethics approval and consent to participate

The study is part of a larger project that has been approved by Helsinki University Hospital ethics review committee (#206/13/03/03/2016 and #303/13/03/01/2011). All samples and data have been derived after written informed consent.

### Consent for publication

Not applicable.

### Availability of data and materials

All unidentifiable data is available within the article and its supporting files. The individual-level sequencing data cannot be made publicly available because the research participant consent does not include authorization to share identifiable data.

### Competing interests

The authors declare that they have no competing interests.

### Funding

Research Council of Finland (#322675, #349760), Sigrid Jusélius Foundation, the Finnish Special Governmental Subsidy for Health Sciences, Research, and Training, the Helsinki University Hospital Comprehensive Cancer Research Funding, Cancer Foundation Finland, and iCAN Digital Precision Cancer Medicine Flagship.

### Authors’ contributions

Study design: OK, EP, UWK; data acquisition: TR, MH, AKL, CH, UWK, OK; PaVaDi pipeline design and implementation: JRK; data analysis and interpretation: JRK, LL, UWK, EP, OK; writing the manuscript JRK, LL, UWK, EP, OK.

All authors have read and approved the final manuscript.

## Acknowledgements

The authors would like to thank the FIMM Genomics unit supported by HiLIFE and Biocenter Finland for genome sequencing and bioinformatics services, and acknowledge CSC – IT Center for Science, Finland, for generous computational resources.

## Notes

### Competing Interest Statement

The authors have declared no competing interest.

### Funding Statement

This study was funded by the Research Council of Finland (322675, 349760), Sigrid Jusélius Foundation, the Finnish Special
Governmental Subsidy for Health Sciences, Research, and Training, the Helsinki University
Hospital Comprehensive Cancer Research Funding, Cancer Foundation Finland, and iCAN
Digital Precision Cancer Medicine Flagship.

### Author Declarations

Ethics committee of Helsinki University Hospital gave ethical approval for this work.

