## Supplementary Text and Supplementary Figures S1-S4 for "Pathogenicity evaluation of coding germline variants identifies rare alleles enriched in hematological patients of a founder population"

Jessica R. Koski[1,2], Laura Langohr[3,2,1,4]*, Tuulia Räisänen[1,2]*, Atte K. Lahtinen[1,2], Marja Hakkarainen[1,2,5], Caroline Heckman[3,4], Ulla Wartiovaara-Kautto[1,2,5]**, Esa Pitkänen[3,1,4]**, Outi Kilpivaara[1,2,4,6,7]**

[1]Applied Tumor Genomics, Research Programs Unit, Faculty of Medicine, University of Helsinki, Helsinki, Finland

[2]Department of Medical and Clinical Genetics, Medicum, Faculty of Medicine, University of Helsinki, Helsinki, Finland

[3]Institute for Molecular Medicine Finland (FIMM), HiLIFE, University of Helsinki, Helsinki, Finland

[4]iCAN Digital Precision Cancer Medicine Flagship, Helsinki, Finland

[5]Department of Hematology, Helsinki University Hospital Comprehensive Cancer Center, University of Helsinki, Helsinki, Finland

[6]HUSLAB Laboratory of Genetics, HUS Diagnostic Center, Helsinki University Hospital, Helsinki, Finland

[7]K. Albin Johansson Cancer Research Fellow, Foundation for the Finnish Cancer Institute, Helsinki, Finland

**PaVaDi implementation of ACMG framework**

**PVS1**

Null variants (nonsense variants, frameshift indels, and canonical splice variants) can often lead to loss of function (LOF). PVS1 was assigned to null variants in LOF intolerant genes, that is, genes that were included in a list of 4,807 LOF intolerant genes, consisting of LOF intolerant genes in ExAC (3,230 genes) and genes harboring at least one LOF variant that is P in ClinVar (1,988 genes) from InterVar (Q. Li & Wang, 2017).

**PS1 and PM5**

If a nucleotide change results in an amino acid that has been previously classified as pathogenic the variant will be given a strong score (PS1). If the nucleotide change results in amino acid change, but has not been classified as pathogenic, it will be given a moderate score (PM5). The list of 38,290 missense variants resulting in pathogenic amino acid change in ClinVar (Landrum et al., 2020) from InterVar (Q. Li & Wang, 2017) was used for scoring.

**BA1, BS1, BS2 and PM2 scoring with population control data**

GnomAD exomes non-cancer was used to score BA1, BS1, BS2 and PM2 criteria. Splice variants were checked using gnomAD genomes. Gene inheritance patterns and age of onset were retrieved from Clinical Genomic Database (CGD) (Solomon et al., 2013). BA1 score is assigned if the variant is present in controls with AF>5%. BS1 score is assigned if variant AF is higher than 0.5%. BS2 score is assigned if the variant is observed in healthy controls and in a gene for which full penetrance is expected at early age (age of onset is ‘pediatric’). Variant allele frequency thresholds are set to AF>1% for autosomal recessive (AR) genes and to AF>0.1% for autosomal dominant (AD) genes. PM2 score will be assigned to variants that are either absent from controls or present at low frequency. If the variant is in an AR or AD/AR gene, the allele frequency threshold is set to AF<1% and to AF<0.1% if it is in an AD or XL gene. For splice variants, the gnomAD genomes threshold for PM2 is AF<0.1%.

PM2 will be updated from moderate to strong (PM2s) if the variant is located in a highly conserved region with conservation score phyloP100way_vertebrate > 7.2.

**PM1**

If the variant is located in a functional domain without benign variants, it will be assigned moderate score PM1. A list of 1,410 domains without benign variants from the InterVar package was used for scoring, including Interpro domains that had only P/LP variants without B/LB or common (MAF>5%) variants.

**PM4 and BP3**

When protein length changes as a result of in-frame deletions or insertions or stop-loss variants in a non-repeat region, it will be assigned moderate strength score PM4. If in-frame deletions or insertions are in a repetitive domain and not in a functional domain, BP3 will be assigned. Repeat regions are retrieved from the UCSC Genome Browser rmsk database (Lee et al., 2022).

**PP2 and BP1**

A missense variant in a gene with low rate of benign missense variation, in which missense variants are a common mechanism of disease, will be assigned score PP2.This includes genes from ClinVar where most of pathogenic variants (>80%, at least one) are missense and <10% of missense variants are benign. A missense variant in a gene in which truncating variants are known to cause disease will be assigned score BP1. This includes genes in which most of the pathogenic variants (>80%) are truncating variants.

A list of 931 genes for PP2 and 1,449 genes for BP1 from the InterVar package were used for scoring.

**PP3 and BP4**

If more than half of the *in silico* tools predicted that the variant has a deleterious effect or has no impact on the gene or gene product, the variant will be assigned PP3 or BP4, respectively. For PP3 and BP4, over 53% out of all the valid predictions need to predict a deleterious effect or no impact, respectively (Kopanos et al., 2019). In the absence of any predictions, if the variant is in a semi-conserved region (phyloP100way_vertebrate > 3.81), PP3 will be assigned, and if the variant is not truncating and not in a conserved region (phyloP100way_vertebrate < 1.4) BP4 will be assigned.

*In silico* tools from dbNSFP4.2a (X. Liu et al., 2020) were used in the analysis. For BayesDel_noAF, DEOGEN2, SIFT, Polyphen2_HDIV, SIFT, Polyphen2_HDIV, PROVEAN,M-CAP, MetaSVM, MutationTaster, MutationAssessor, MetaRNN, LIST-S2, PrimateAI we used the predictions for deleterious (D) and tolerated (T). For REVEL, CADD, FATHMM_MKL, MVP, DANN we used thresholds 0.5, 15, 0.5, 0.75, 0.93 respectively.

**PP5 and BP6**

If a variant is reported as pathogenic or benign in ClinVar (Landrum et al., 2020) without strong evidence, it is assigned supporting PP5 or BP6, respectively. If a variant has strong ClinVar evidence, or CLNREVSTAT is “multiple submitters and no conflicts", “reviewed by expert panel” or “practice guideline”, the labels strong PP5_s and BP6_s are used instead.

**BP7**

A synonymous variant will be assigned BP7 score if it has no effect on splicing and is not at a highly conserved locus. Effect on splicing is predicted with random forest (RF) and adaptive boosting (ADA) scores from dbNSFP4.2a (X. Liu et al., 2020) with values below 0.515 and 0.708, respectively, considered to have no impact on splicing.

#### Evaluating the effect of case-control imbalance to variant enrichment

We observed the minor allele frequency (mAF) in controls to explain $R^2$=47% of variability in variant enrichment mAF(case)/mAF(control) in the set of 156,245 variants discovered in our study set. This correlation is due to rare variants: in the set of variants with mAF(control)>0.001, this effect is minimal ($R^2$=1.4%). To evaluate how much of the variability is due to the imbalance in sizes of case and control sets, we performed a simulation experiment of 10,000 variants. For each simulated variant, mAF(control) was assigned to be mAF(gnomAD Finns non-cancer) of a randomly selected variant in the study set. Then, genotypes for 511 cases were randomly determined with mAF=mAF(control) (*i.e.*, case genotypes were drawn from the distribution of control genotypes), yielding mAF(case) for each variant. In the simulated dataset, we observed a similar skew as in our study set, albeit of smaller magnitude ($R^2$=16%; **Suppl. Fig. S1**). We thus conclude that about a third (16/47=34%) of the observed shared variability is due to the case-control imbalance.

#### Biallelic events

Somatic second hits and LOH may give further evidence for variant pathogenicity (Huang et al. 2018). To detect potential biallelic events in genes carrying a P/LP germline variant, we studied LOH and biallelic events in tumor samples that were available for 460 patients. We identified secondary somatic mutations in five genes where a P/LP germline variant was coupled with a missense or truncating somatic mutation in the same gene (Supplementary Table 14). Two Finnish founder variant carriers *FANCM* Gln1701Ter and *ATM* Ala2524Pro (Pylkäs et al., 2007) were found with secondary somatic mutation with tumor allelic fractions 3.5% and 15.6%, respectively. In addition, another patient carrying the same *ATM* mutation showed LOH (FDR=0.0024). *ATM* Ala2524Pro has earlier been reported as a high-risk allele for breast cancer in Finland (Kankuri-Tammilehto et al., 2023). Other genes with identified somatic secondary mutations were *LPIN2* and *DDX41* with allelic fractions 2% and 23%, respectively. Somatic findings act as further evidence on variant pathogenicity on HMs for these genes.

### Supplementary Figures

**Supplementary Figure S1**. A scatterplot of mAF(control) (X-axis; logarithmic scale) versus mAF(case) (Y-axis; logarithmic scale with added coordinate noise) of 10,000 variants in a simulated set of 511 individuals where case genotypes are randomly assigned with mAF=mAF(control).

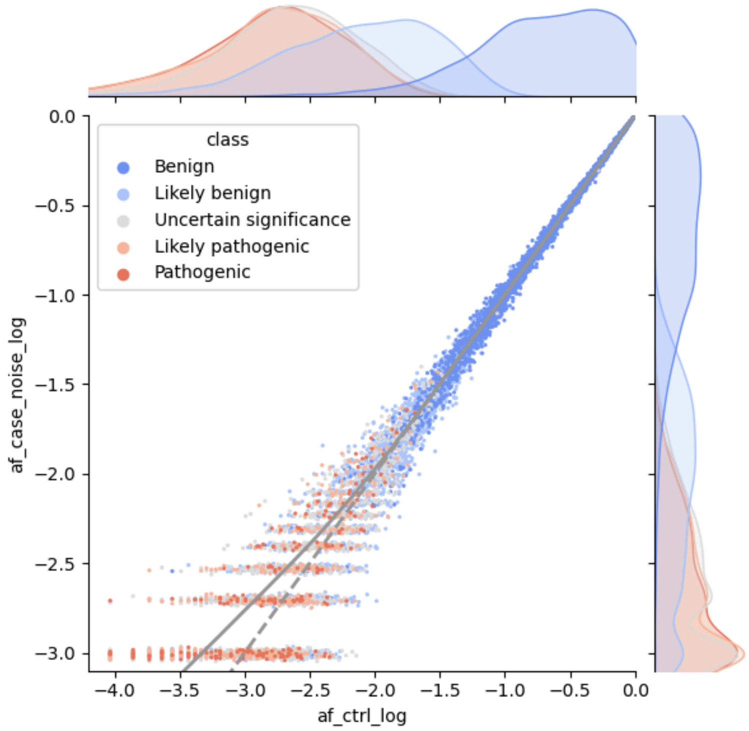

**Supplementary Figure S2.** Graphical representation of pathogenicity classification. Variants will be assigned to a pathogenicity class based on the evidence filling the criteria. When not enough criteria is filled to reach P/LP or B/LB classification, the variant will be assigned VUS. If both P/LP and B/LB classification is annotated, the conflicting criteria ends in VUS classification.

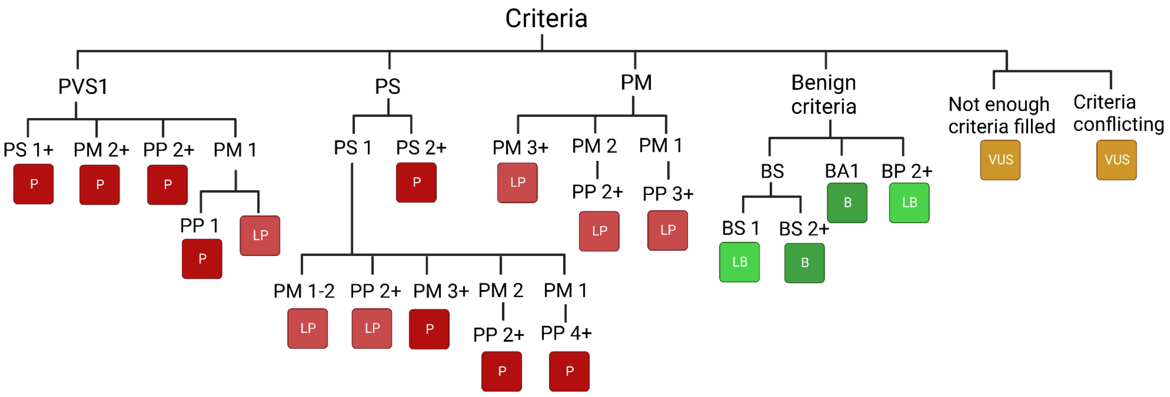

**Supplementary Figure S3.** Burden test histogram for unadjusted p-values.

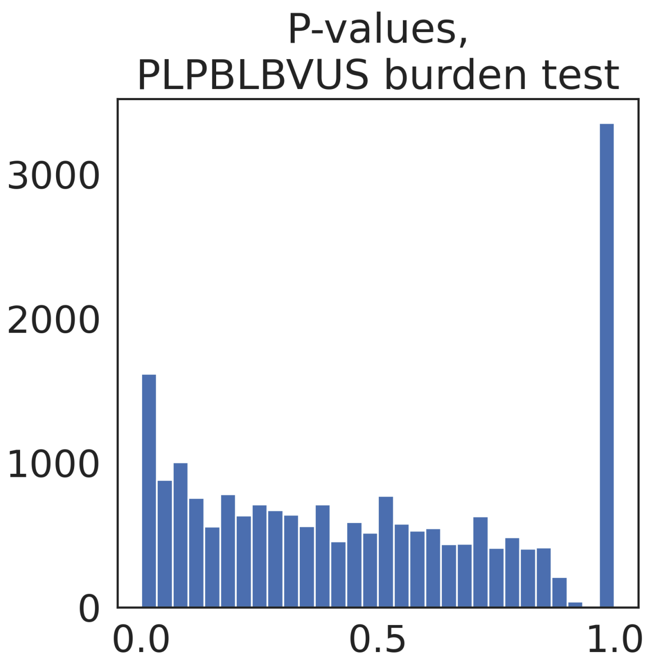


**Supplementary Figure S4.** Burden test QQ-plot unadjusted p-values.

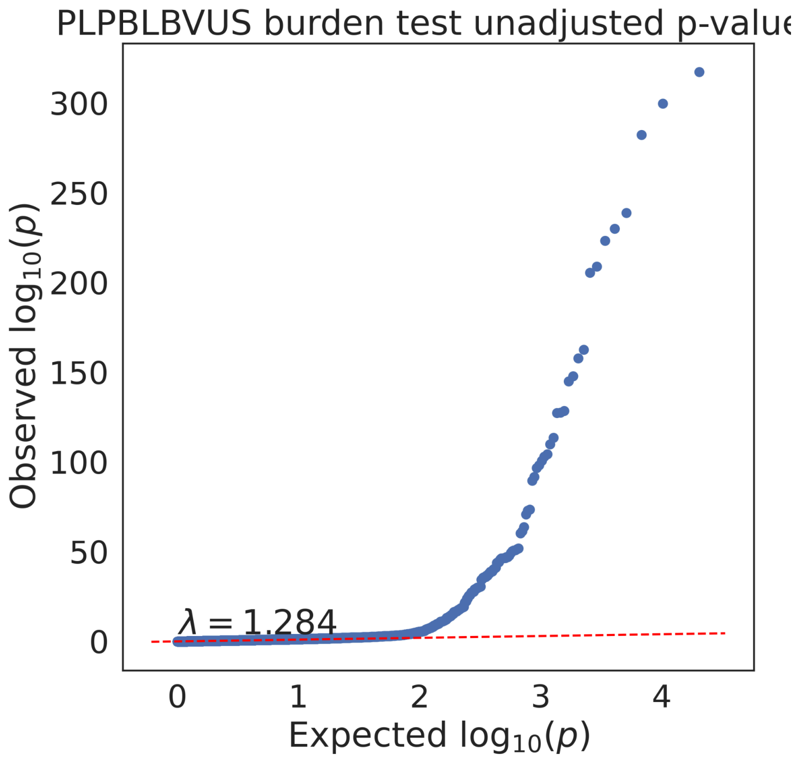
